# Burden of Chronic Kidney Disease in China, 1990-2021: Findings from the 2021 Global Burden of Disease Study

**DOI:** 10.64898/2026.06.06.26355056

**Authors:** Mengwei Wang, Taoran Zhao, Heng Wang, Shulin Hou, Yongqiang Fu

## Abstract

**Introduction:** To investigate the epidemiological characteristics of chronic kidney diseases (CKD) in China in 2021 and its trends between 1990 and 2021, in the context of significant population growth and lifestyle changes over the past 30 years that have likely influenced the CKD spectrum.

**Methods:** Data on CKD prevalence, mortality, disability-adjusted life-years (DALY), and risk factors were obtained from the Global Burden of Disease Study 2021. The estimated decadal percentage changes were calculated to evaluate changes in trends in prevalence, mortality and disease burden.

**Results:** In 2021, an estimated 118.4 (95% UI 109.4 to 127.5) million people in China were affected by CKD, contributing to 204 230 (95% UI 164 736 to 246 372) deaths and 6.13 (95% UI 5.18 to 7.21) million DALY. Although CKD due to diabetes mellitus and hypertension accounted for less than a quarter of all cases, they were responsible for over 90% of CKD-related deaths. Over the past three decades, CKD mortality and DALY rates have steadily increased, although the prevalence has stabilized in the last decade. Diabetes mellitus type 2 and hypertension have emerged as key drivers of CKD burden in China.

**Conclusions:** The CKD burden in China shows a dual pattern of rising incidence and high mortality from diabetes and hypertension-related chronic kidney disease, alongside persistently high years lived with disability from glomerulonephritis and other causes.

## Introduction

It is estimated that over 850 million individuals worldwide are affected by kidney diseases, including acute kidney injury, chronic kidney disease (CKD), and treated kidney failure[1]. By 2040, CKD is projected to become the fifth leading cause of death globally, and in countries with higher life expectancy, it is anticipated to rank as the second leading cause of death by the end of the century[2]. China is one of the countries with the highest number of patients with CKD, accounting for one-fifth of the global CKD population in 2019[3]. According to a nationwide survey conducted in China in 2018-2019, the estimated prevalence of CKD was 8.2% in the national population[4].

As a comprehensive study of global health loss, the Global Burden of Diseases, Injuries, and Risk Factors Study (GBD) 2021 provides current information on the distribution and burden of diseases and injuries across time, age, sex, location, and sociodemographic group[5]. In the present study, we analyzed the prevalence, mortality, and disability-adjusted life year (DALY) of CKD in the Chinese population by age and sex for the period 1990-2021 using the results from GBD 2021, aiming to provide a better understanding of the disease burden and trends of CKD at the national, age, and sex levels.

In the past three decades, along with economic growth and rapid urbanization, China has experienced dramatic societal and environmental changes, and the spectrum of CKD in China has shifted[6]. Data from the Hospital Quality Monitoring System (HQMS) in China revealed that glomerulonephritis was the leading cause of CKD in China until 2011. Since then, the proportion of CKD due to diabetes mellitus first exceeded that due to glomerulonephritis[7]. An increasing trend in CKD due to hypertension was also revealed via HQMS data from 2010 to 2015[8]. In 2016, diabetes (25.7%) and hypertensive nephropathy (21.4%) became the two most common causes of CKD among hospitalized patients[9]. In this study, we not only present the latest epidemiological and disease burden data on CKD caused by diabetes and hypertension in China from GBD 2021 but also analyze and visualize the proportion of the subsets of CKD within the overall CKD burden in China and their trends over the past 30 years. In addition, we propose constructive prevention strategies to address this growing public health issue.

## Methods

### Data sources

The GBD 2021 examined 371 diseases and injuries, as well as 84 risk factors, by age and sex globally across 204 countries and regions, covering the years from 1990 to 2021[5]. In the GBD study, all appropriate data and statistical methods were employed to leverage predictive covariates and geographical proximity to countries with available data, thereby generating estimates of incidence, prevalence, deaths, and DALYs for all regions. We extracted data on the burden of CKD in China from 1990 to 2021 from the GBD 2021 study and analyzed the burden of CKD in China in 2021 as well as the trends from 1990 to 2021.

### DALYs, years of life lost (YLLs) and years lived with disability (YLDs)

Estimates of deaths due to CKD were multiplied by the standard life expectancy at each age to calculate the YLLs for CKD[5]. Non-fatal CKD cases were categorized into 15 distinct sequelae based on the CKD stage and anemia severity[5]. The YLDs for each sequela was then determined by multiplying their prevalence by the associated disability weight[5]. DALYs were defined as the sum of YLLs and YLDs, representing one lost year of a healthy life[5].

### Definition of CKD

CKD in GDB was defined as an abnormality of kidney function, indicated by an estimated glomerular filtration rate (eGFR) based on serum creatinine measurements lower than 60 mL/min/1.73 m^2^ and/or a urine albumin to-creatinine ratio (ACR) higher than 30 mg/g [(Supplementary data, Table S1), also published elswhere[5]].

### Causes of CKD

In the GDB 2021, the burden of CKD was estimated for each of the five causes: type 1 diabetes, type 2 diabetes, glomerulonephritis, hypertension, and a residual category of other and unspecified causes. For every individual with CKD, the International Classification of Diseases and Injuries (ICD) codes for primary renal diseases were used to map individuals to GBD cause groupings (Supplementary data, Table S2). Individuals with CKD but no ICD code for primary renal disease were classified as having CKD of an uncertain cause. In this study, we focused on CKD caused by diabetes and hypertension; therefore, all other causes were combined into a single category labeled as glomerulonephritis and other causes.

### Study population

Patient age was classified into 17 subgroups: ages 0-14, ages 15-19, ages 20-24, ages 25-29, ages 30-34, ages 35-39, ages 40-44, ages 45-49, ages 50-54, ages 55-59, ages 60-64, ages 65-69, ages 70-74, ages 75-79, ages 80-84, and > 85 years.

### Estimation of risk factor

GBD 2021 includes 70 risk factors, categorized into the three groups shown above: environmental/occupational risks, behavioral risks, and metabolic risks. We extracted the risk factors contributing to the CKD-related DALYs and their corresponding contribution levels. In the GBD study, CKD was considered both a disease and a metabolic risk factor. In this analysis, we considered CKD as a disease. In addition, 14 other risk factors contribute to the burden of CKD in China.

### Statistical analysis

To account for uncertainty in primary data sources, data manipulations, measurement error and model selection, all entities quantified in the 2021 GBD study were estimated 500 times in the ensemble and meta-regression models, resulting in final estimates with 95% uncertainty intervals (UI), represented by the 2.5th and 97.5th percentile values of the 500 draws. The rates in our projections are displayed per 100 000 people. The age-standardized prevalence rate, age-standardized mortality rate and age-standardized DALYs rates were used to eliminate the effect of differences in age distribution. All statistical analyses were conducted using the R Studio software (version 1.4.1106) and GraphPad Prism (version 8.0).

## Results

### CKD Burden in China in 2021

#### Prevalence

In China, it is estimated 118.4 million (95% UI 109.4 to 127.5) prevalent CKD cases in 2021, accounting for 8.32% (95% UI 7.69 to 8.96) of the national population (Table 1, Fig. 1. a). This figure represents 17.6% of the global CKD burden. The prevalence of CKD was slightly higher in females than in males (Fig. 1. b and 1. c), with an age-standardized prevalence rate of 6 710 (95% UI 6 230 to 7 235) per 100 000 population for females, as opposed to 5 793 (95% UI 5 390 to 6 225) per 100 000 population for males (Table 1).

**Fig. 1.**
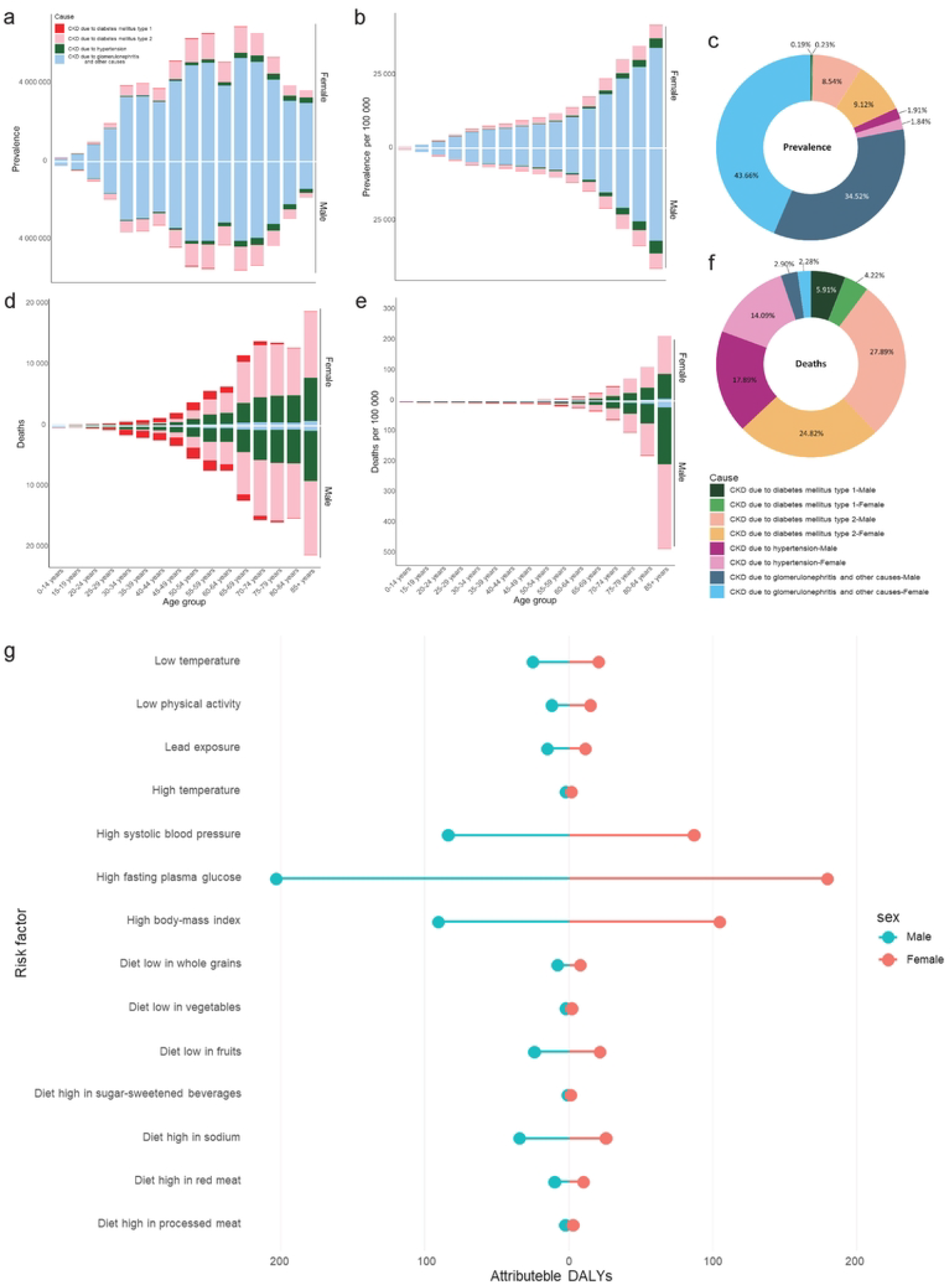
Prevalence and deaths of CKD in China by age, sex and causal attribution in 2021. **a** Prevalence cases. **b** Prevalence rate. **c** Proportion of prevalence cases. **d** Deaths cases. **e** Deaths rates. **f** Proportion of deaths cases. **g** Attributable DALY of CKD in China by risk factors and sex in 2021.

#### Deaths

In 2021, CKD resulted in an estimated 204 230 (95% UI 164 736 to 246 372) deaths in China, with an age-standardized death rate of 10 per 100 000 population (95% UI 8 to 12), comprising 1.7% of national deaths (Table 1, Fig. 1. d). A higher mortality rate among males than females was observed in patents with CKD. Of these deaths, 111 482 (95% UI 82 929 to 144 064) were male, with an age-standardized death rate of 13 per 100 000 population (95% UI 10 to 17), while 92 747 (95% UI 72 663 to 117 085) were female, with an age-standardized death rate of 8 per 100 000 population (95% UI 6 to 11) (Table 1) (Fig. 1. e and 1. f).

#### DALYs, YLDs and YLLs

In 2021, CKD included in this analysis caused 1.69 million (95% UI 1.20 to 2.20) YLDs and 4.44 million (95% UI 3.57 to 5.39) YLLs (Table2), contributing to a total of 6.13 million (95% UI 5.18 to 7.21) DALYs (Table 1, Fig. 2. a, 2. d and 2. g). The burden due to CKD was observed across all age groups, emerging before the age of 20 years in individuals with CKD due to diabetes mellitus type 1, glomerulonephritis and other causes, and continuing into older ages in those with CKD due to diabetes mellitus type 2 and hypertension. While the relative contribution of each disorder varied by age and sex, the number of YLLs increased from childhood and adolescence, peaked between 65 and 74 years, and decreased after the age of 75 years (Fig. 2. g). From a gender perspective, the overall YLLs due to CKD from various causes were higher in males than in females (Fig. 2. h and 2. i). Figure 2 shows the national CKD DALYs, YLDs and YLLs by cause, age, and sex in 2021.

**Fig. 2.**
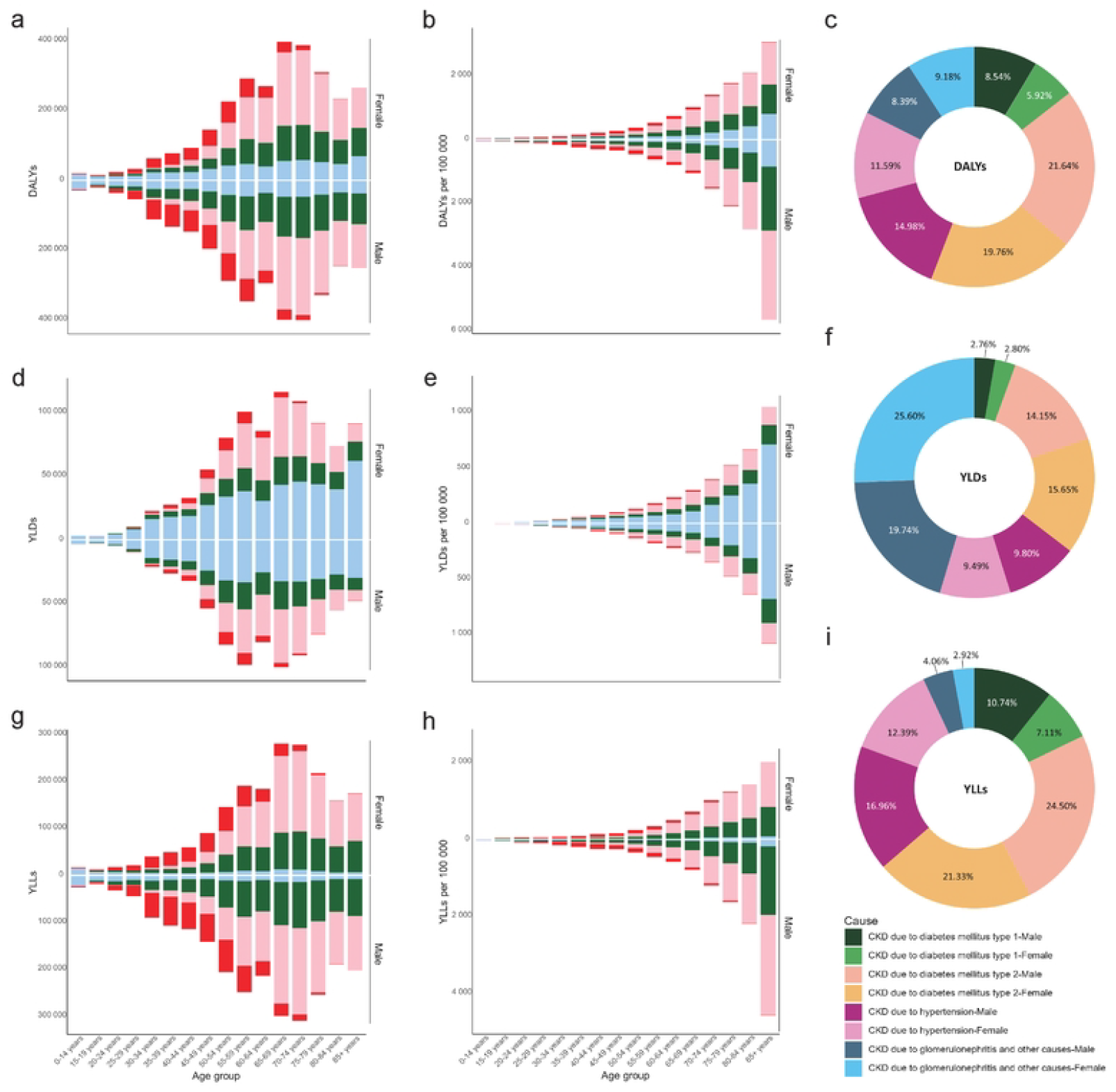
DALYs, YLDs and YLLs of CKD in China by age, sex and causal attribution in 2021. **a** DALYs cases. **b** DALYs rate. **c** Proportion of DALYs cases. **d** YLDs cases. **e** YLDs rates. **f** Proportion of YLDs cases. **g** YLLs cases. **h** YLLs rates. **i** Proportion of YLLs cases. Abbreviations: CKD, chronic kidney disease; YLDs, years of life lost due to premature mortality; YLLs, years lived with disability.

### Causal attribution

#### CKD due to diabetes mellitus

In 2021, the prevalence of CKD due to both types of diabetes mellitus in China was 21.4 million (95% UI 19.6 to 23.2). The age-standardized prevalence rate of CKD due to diabetes mellitus type 2 was 1 053 (95% UI 971 to 1 139) per 100 000 population, with an age-standardized mortality rate of 5 (95% UI 4 to 6) per 100 000 population. The age-standardized prevalence rate of CKD due to diabetes mellitus type 1 was 28 (95% UI 23 to 33) per 100 000 population, and the age-standardized mortality rate was 1 (95% UI 0 to 1) per 100 000 population (Table 1). Although CKD due to diabetes accounts for only 18.08% of the total CKD prevalence (Fig. 1. c), it is responsible for 62.84% of the total CKD-related deaths (Fig. 1. f). Additionally, CKD due to diabetes accounted for 55.86% of CKD-related DALYs (Fig. 2. c), including 35.36% of CKD YLDs (Fig. 2. f), and 63.68% of CKD YLLs (Fig. 2. i). While women have higher prevalence rates (Fig. 1. b) and YLDs rates (Fig. 2. e), men experience higher mortality rates (Fig. 1. e) and YLLs rates (Fig. 2. h), particularly among those aged 75 years and older.

#### CKD due to hypertension

In 2021, in China, there were 4.4 million (95% UI 4.1 to 4.8) cases of CKD attributed to hypertension. The age-standardized prevalence rate of CKD due to hypertension was 223 (95% UI 206 to 244) per 100 000 population, and the age-standardized mortality rate was 3 (95% UI 2 to 4) per 100 000 population (Table 1). CKD due to hypertension accounted for 3.75% of the total CKD cases in China (Fig. 1. c); however, it contributed to 31.98% of CKD-related deaths (Fig. 1. f) and 26.57% of CKD-related DALYs (Fig. 2. c). It was responsible for 19.29% of the total CKD YLDs (Fig. 2. f) and 29.35% of the total CKD YLLs (Fig. 2. i). Compared to females, males with CKD due to hypertension exhibited higher mortality rates (Fig. 1. e) and YLLs rates (Fig. 2. h).

#### CKD due to glomerulonephritis and other causes

In China, there were 92.6 million (95% UI 85.5 to 99.9) cases of CKD due to glomerulonephritis and other causes, accounting for 78.18% of all CKD patients in the country (Fig. 1. c). The age-standardized prevalence rate of CKD due to glomerulonephritis and other causes was 4 943 (95% UI 4 578 to 5 342) per 100 000 population (Table 1). In 2021, CKD due to glomerulonephritis and other causes caused 10 585 (95% UI 7 605 to 14 222) deaths in China, representing 5.18% of CKD-related deaths (Fig. 1. f). Despite its low mortality rate, the burden of CKD due to glomerulonephritis and other causes remains significant. In 2021, YLDs attributed to this cause accounted for 45.34% of the total CKD YLDs (Fig. 2. f), and the DALYs were 1,076,654 (95% UI 823 677 to 1 359 530), representing 17.57% of the total CKD DALYs (Fig. 2. c).

#### Risk factors

Three metabolic risks, including high fasting plasma glucose, high body-mass index (BMI) and high systolic blood pressure were the leading risk factors for CKD DAYLs in both sexes in 2021 (Fig. 1. g). Among these factors, high fasting plasma glucose shows higher attributable DALYs in males than in females, while high BMI shows higher attributable DALYs in females than in males. Factors such as a diet low in whole grains, vegetables, and fruits, and a diet high in sugar-sweetened beverages, sodium, red meat, and processed meat have relatively minor but still noteworthy contributions to CKD DALYs. In most cases, these dietary factors show slightly higher attributable DALYs for males than for females. Environmental factors, particularly low temperatures and lead exposure, also have a noticeable impact on CKD DALYs. The chart indicates that these factors contributed slightly more to the disease burden in males compared to females. The chart also shows that low physical activity has a greater impact on CKD DALYs in females compared to males.

#### Temporal trends of CKD since 1990 in China Prevalence

The past decade has witnessed the most rapid increase in CKD prevalence rate (Fig. 3. a). Specifically, the prevalence count increased by 10.2% (95% UI 8.8% to 11.7%), with a corresponding increase in the prevalence rate of 3.0% (95% UI 1.6% to 4.4%) from 1990 to 2000. Between 2000 and 2010, there was a further 24.6% (95% UI 22.5% to 27.1%) increase in the prevalence count, with the prevalence rate rising by 17.4% (95% UI 15.3% to 19.7%). The prevalence count and rate increased by 27.9% (95% UI 25.5% to 30.2%) and 20.2% (95% UI 17.9% to 22.4%), respectively, from 2010 to 2021 (Table 3).

**Fig. 3.**
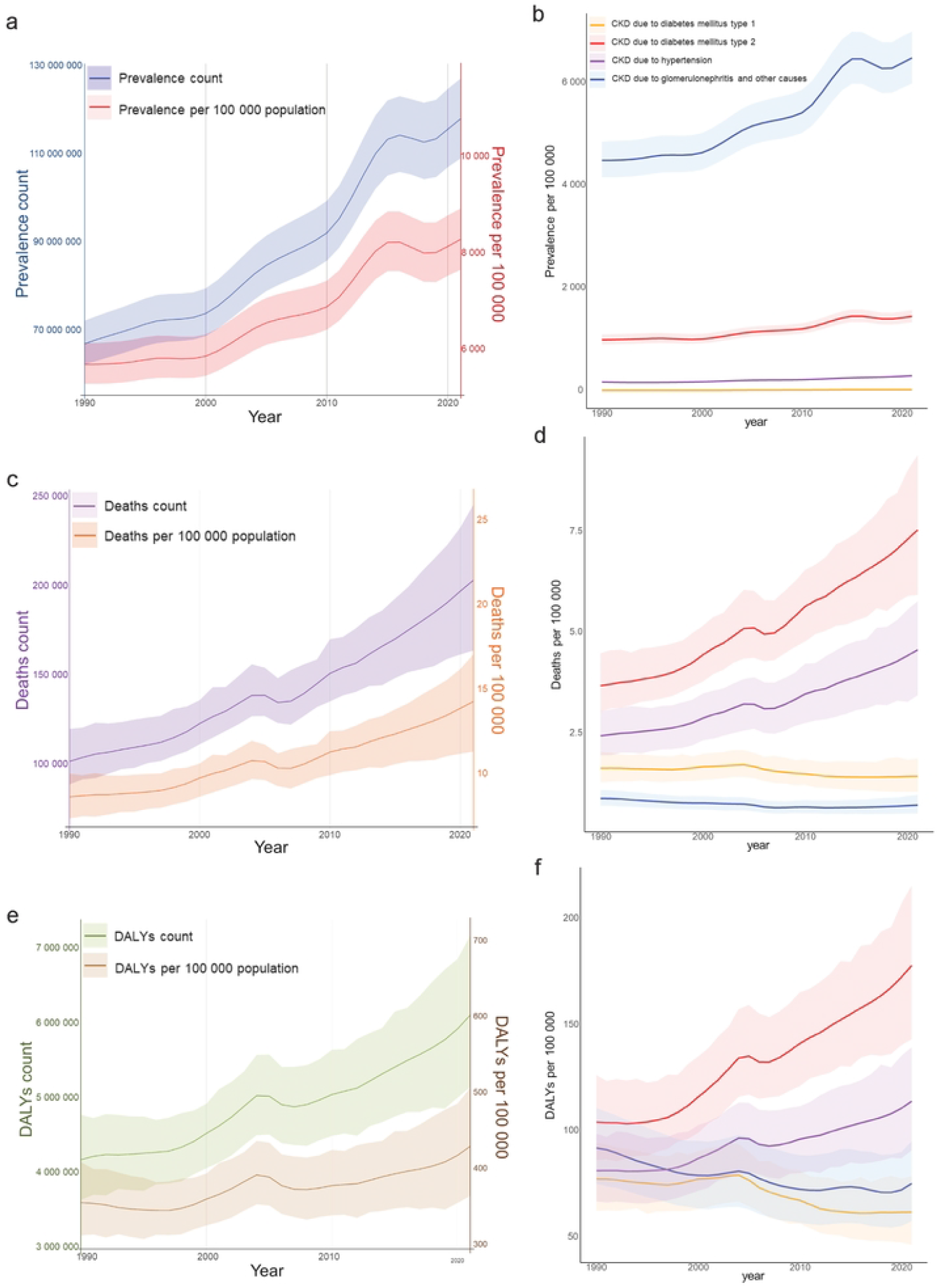
Changes in CKD burden in China from 1990 to 2021. **a** prevalence cases and prevalence rates. **b** different attribution prevalence rates. **c** deaths cases and deaths rates. **d** different attribution mortality rates. **e** DALYs cases and DALYs rates. **f** Different attribution DALYs rates.

Since 1990, in China, CKD due to diabetes mellitus type 1 has had a stable and the lowest prevalence rate in China. The prevalence rate of CKD due to hypertension has been slightly higher than that due to diabetes mellitus type 1, with a continuous rise in trends since 2000. The prevalence rate of CKD resulting from diabetes mellitus type 2 has shown a continuously increasing trend over the past two decades, with a particularly rapid rise between 2010 and 2015, followed by a gradual deceleration in the upward trend. Lastly, the trend of prevalence rate of CKD due to glomerulonephritis and other causes, which constitute the majority of the CKD population in China, mirrors the overall trend in CKD prevalence rate over the past 30 years (Fig. 3. b).

#### Deaths

Since 1990, both the number of deaths and the mortality rate associated with CKD in China have exhibited a continuous and accelerating increase, although this rise has been relatively slower compared to the growth in prevalence rates. Between 1990 and 2000, the number of deaths increased by 20.7% (95% UI 4.6% to 38.6%), with the mortality rate rising by 12.8% (95% UI -2.2% to 29.5%). From 2000 to 2010, the number of deaths further increased by 22.7% (95% UI 10.6% to 36.5%), and the mortality rate increased by 15.5% (95% UI 4.2% to 28.6%). The trend continued from 2010 to 2021, with a 34.3% (95% UI 9.6% to 62.3%) increase in the number of deaths and a 26.2% (95% UI 3.0% to 52.5%) increase in the mortality rate (Table 2) (Fig. 3. c).

CKD due to diabetes mellitus type 2 or hypertension accounts for the majority of CKD-related deaths, and mortality rates associated with these two causes have also been continuously rising. In contrast, the mortality rate of CKD due to diabetes mellitus type 1 has gradually declined since 2005. The death rate from CKD due to glomerulonephritis and other causes has remained consistently low over the past 30 years (Fig. 3. d).

#### DALYs

In China, when considering all subtypes combined, the burden of CKD experienced a slight increase between 1990 and 2000, with the DALYs count of CKD rising by 8.4% (95% UI -2.7% to 20.8%), and the DALYs rate per 100 000 increasing by only 1.3% (95% UI -9.0% to 12.9%). From 2000 to 2010, despite some fluctuations, the CKD DALYs count increased by 11.4% (95% UI 2.1% to 21.8%), and the DALYs rate per 100 000 increased by 4.9% (95% UI -3.8% to 14.7%). However, from 2010 to 2021, the CKD burden increased sharply, with the CKD DALYs count rising by 20.9% (95% UI 3.8% to 40.1%), and the DALYs rate per 100 000 increased by 13.6% (95% UI -2.5% to 31.6%) (Table 3) (Fig. 3. e).

Over the past 30 years, the DALYs rate per 100 000 with CKD due to diabetes mellitus type 2 or hypertension has shown slight fluctuations but an overall steady increase, with a notable acceleration in the upward trend after 2000. In contrast, since 1990, the DALYs rate per 100 000 for CKD due to type 1 diabetes has remained at its lowest level, with minimal change over the 30-year period, indicating that the burden of CKD related to this cause has remained relatively low. Similarly, CKD due to glomerulonephritis and other causes showed a relatively low DALYs rate, with an overall downward trend throughout the study period (Fig. 3. f).

## Discussion

In the past two years, several epidemiological studies on CKD in China have been conducted, including analyses based on GBD 2019 data [3, 10] and report by the sixth China Chronic Disease and Risk Factor Surveillance (CCDRFS) [4]. However, these studies either focused on specific disease subtypes [10] or were based on data collected before 2020. This study provides a comprehensive national picture of CKD in China in 2021. This study highlights the significant burden caused by a dual pattern of rising incidence and high mortality from diabetes and hypertension-related chronic kidney disease, along with persistently high years lived with disability from glomerulonephritis and other causes.

In the 2018-2019 CKD survey conducted by the sixth CCDRFS, the number of CKD cases among individuals aged > 18 years was reported to be 82 million, with a prevalence rate of 8.2% (95% CI, 7.8%-8.6%) [4]. According to GBD 2021 data, the corresponding figures for all age groups were 113.9 million (95% UI, 105.2 to 122.7) and 8.1% (95% UI, 7.4%-8.7%). Considering the very low incidence of CKD among individuals under 18 years of age, the CKD prevalence data estimated by the GBD 2021 statistical method may be close to, but slightly higher than, the results obtained using the CCDRFS survey method. The low awareness among patients with early-stage CKD regarding about their condition has also made it challenging to accurately assess the prevalence of CKD. According to data from the sixth CCDRFS, the weighted awareness of CKD in the Chinese population was only 10% in 2018-2019 [4]. The data used in the GBD 2021 study were based on estimates, which may introduce potential bias or imprecision. Although efforts have been made to correct for underreporting and measurement errors, gaps in healthcare records, particularly among underdiagnosed populations, could impact the accuracy of estimates [5]. Nevertheless, in addition to prevalence data, the GBD 2021 statistics also provide mortality data and DALYs figures, reflecting the burden of disease.

Therefore, these findings based on the GDB 2021 remain valuable for informing decisions related to disease prevention and control.

While CKD prevalence and death rates increase with age, the burden was most significant among individuals aged 65-74, peaking before gradually decreasing after age 75 years. The ageing of China’s population and increased life expectancy may explain this pattern. According to China’s most recent census, the number of people aged over 65 years reached 190.64 million in 2020, accounting for 13.50% [11]. Additionally, the life expectancy in China rose from 68.6 years in 1990 to 78.2 years in 2021, reflecting broader demographic shifts that likely contribute to the rising burden of CKD in older adults [12].

The findings from the GBD 2021 demonstrate that in 2021, CKD due to type 2 diabetes mellitus accounted for 18% of all CKD cases; however this group was responsible for 62.84% of CKD-related deaths and 55.86% of CKD-related DALYs. CKD due to hypertension contributed to 3.75% of cases but also accounted for 31.98% of CKD-related deaths and 26.57% of CKD-related DALYs. These findings indicate that while these two conditions are not the most common causes of CKD, they lead to significantly more severe disease progression and outcomes compared to glomerulonephritis and other causes. This also explains why statistics from hospitalized patients show that CKD caused by diabetes and hypertension accounts for the highest proportion [6, 13].

The DALYs rate due to CKD caused by diabetes mellitus type 2 or hypertension has shown particularly sharp increases in the past decade, aligning with the growing prevalence of these conditions in the Chinese population. The prevalence of diabetes among Chinese adults increased from 10.9% (95% CI, 10.4%-11.5%) in 2013 to 12.4% (95% CI, 11.8%-13.0%) in 2018 (P < .001) [14]. Similarly, the estimated prevalence of prediabetes rose from 35.7% (95% CI, 34.2%-37.3%) in 2013 to 38.1% (95% CI, 36.4%-39.7%) in 2018 (P = .07) [14]. Despite improvements in diabetes management and treatment, the prevalence of kidney disease among individuals with diabetes has remained steady at approximately 35% [15]. The standardized prevalence of hypertension among adults aged 18-69 in China increased from 20.8% (95% CI, 19.0%-22.5%) in 2004 to 29.6% (95% CI, 27.8%-31.3%) in 2010, followed by a decline to 24.7% (95% CI, 23.2%-26.1%) in 2018 [16]. The relationship between blood pressure and CKD is complex, as hypertension can contribute to and result from declining kidney function [17]. According to the Sixth CCDRFS, 60% of Chinese adults with CKD also have hypertension [4]. It is noteworthy that the impact of diabetes and hypertension on CKD varies across different sexes and age groups, with males generally exhibiting higher CKD-related mortality and YLLs compared to females, particularly in individuals aged 75 years and older.

The dramatic increase in CKD attributable to diabetes and hypertension reflects changes in lifestyle and metabolic risk factors. Specifically, high fasting plasma glucose level, high BMI, and high systolic blood pressure were identified as the leading metabolic risk factors contributing to CKD DALYs in both men and women. In addition, the GBD 2021 data highlight that while metabolic factors are the dominant contributors, these behavioral and environmental risks, particularly in the male population, further exacerbate CKD progression.

Compared with individuals without CKD, those with CKD exhibit a higher prevalence of cardiovascular disease and poorer cardiovascular outcomes [18]. Additionally, CKD is associated with cognitive impairment, not only in the elderly[19, 20], but also among individuals aged 20-59 with moderate CKD[21]. Effective prevention and management of CKD are crucial for the nation’s overall health management strategy.

## Conclusion

In summary, the findings from the GBD 2021 study highlight the rapidly increasing burden of CKD in China from 1990 to 2021, largely driven by diabetes and hypertension. Mitigating metabolic risk factors through focused lifestyle interventions, enhanced screening, and early detection, especially in high-risk groups, will be essential for reducing future disease burden.

## Data Availability

No data were generated by this study. The data used in this study are publicly available from the Global Burden of Disease Study 2021 via the Global Health Data Exchange (GHDx) at: https://vizhub.healthdata.org/gbd-results/.

## Statements

### Ethics Approval

The Global Burden of Disease Study 2021 was approved by the University of Washington Institutional Review Board. All original data collection complied with the Declaration of Helsinki.

## Consent to Participate

Not applicable. This study used only anonymized, aggregated public data from the GBD 2021, which contains no personally identifiable information; therefore, individual informed consent was not required.

## Author Contributions

Mengwei Wang and Yongqiang Fu conceived and designed the study and had full access to all data in the study. Taoran Zhao and Yongqiang Fu analyzed the data and drafted the manuscript. Mengwei Wang, Shulin Hou and Heng Wang contributed to the data acquisition and data visualization.

## Data Availability Statement

The data used in this study are publicly available through the Global Health Data Exchange (https://vizhub.healthdata.org/gbd-results/).

